# Deep reinforcement learning framework for controlling infectious disease outbreaks in the context of multi-jurisdictions

**DOI:** 10.1101/2022.10.18.22281063

**Authors:** Seyedeh Nazanin Khatami, Chaitra Gopalappa

## Abstract

In the absence of pharmaceutical interventions, social distancing and lockdown have been key options for controlling new or reemerging respiratory infectious disease outbreaks. The timely implementation of these interventions is vital for effectively controlling and safeguarding the economy.

Motivated by the COVID-19 pandemic, we evaluated whether, when, and to what level lockdowns are necessary to minimize epidemic and economic burdens of new disease outbreaks. We formulated the question as a sequential decision-making Markov Decision Process and solved it using deep Q-network algorithm. We evaluated the question under two objective functions: a 2-objective function to minimize economic burden and hospital capacity violations, suitable for diseases with severe health risks but with minimal death, and a 3-objective function that additionally minimizes the number of deaths, suitable for diseases that have high risk of mortality. A key feature of the model is that we evaluated the above questions in the context of two-geographical jurisdictions that interact through travel but make autonomous and independent decisions, evaluating under cross-jurisdictional cooperation and non-cooperation.

In the 2-objective function under cross-jurisdictional cooperation, the optimal policy was to aim for shutdowns at 50% and 25% per day. Though this policy avoided hospital capacity violations, the shutdowns extended until a large proportion of the population reached herd immunity. Delays in initiating this optimal policy or non-cooperation from an outside jurisdiction required shutdowns at a higher level of 75% per day, thus adding to economic burdens. In the 3-objective function, the optimal policy under cross-jurisdictional cooperation was to aim for shutdowns of up to 75% per day to prevent deaths by reducing infected cases. This optimal policy continued for the entire duration of the simulation, suggesting that, until pharmaceutical interventions such as treatment or vaccines become available, contact reductions through physical distancing would be necessary to minimize deaths. Deviating from this policy increased the number of shutdowns and led to several deaths.

In summary, we present a decision-analytic methodology for identifying optimal lockdown strategy under the context of interactions between jurisdictions that make autonomous and independent decisions. The numerical analysis outcomes are intuitive and, as expected, serve as proof of the feasibility of such a model.

## 1. Introduction

Timely implementations of pharmaceutical and non-pharmaceutical interventions (NPI) are critical for effective control of new infectious disease outbreaks. Delay in response causes enormous disease and economic burdens, as seen during the COVID-19 outbreak caused by the SARS-Cov2 virus [1].

In the event of new respiratory infectious disease outbreaks, when pharmaceutical interventions are unavailable, NPIs are the only options, as was the case with COVID-19. Effective NPI options include facemask-use and social distancing [2]. Social distancing could include physical distancing (e.g., by 3ft or 6ft) or partial lockdowns. While facemasks and physical distancing could be the most economically feasible options, lockdowns may be necessary for highly contagious viruses such as the SARS-Cov2. While locking-down early in the pandemic would be suitable for reducing disease burden, it may unnecessarily add to the economic burden. On the other hand, delaying the lockdown or improper phasing of lockdowns can significantly amplify both economic and disease burdens [3].

In this context, through timely implementation of lockdowns, governmental public health agencies play a key role in effective containment of new outbreaks. Furthermore, though public health decisions are autonomous to each jurisdiction, e.g., in the United States, local COVID-19 prevention guidelines were determined by individual states [4], the epidemic can be influenced by outside jurisdictions through travel.

A methodology that can help determine whether and when a lockdown is necessary, to what level, and how to phase out a lockdown would be a critical part of a pandemic preparedness plan. While surveillance systems to help identify new outbreaks would be a crucial part of this preparedness plan, because of the delay in diagnosis of cases, informing decisions only based on data collected through these systems will not be sufficient. Surveillance data combined with epidemic projections through the use of dynamic mathematical models can help identify optimal control policies, including whether a partial shutdown will be necessary [4, 5]. In this study, we formulated the question of whether and when a lockdown is necessary, to what level, and how to phase out a lockdown as a sequential decision-making problem using Markov decision process (MDP) and solved using Deep Q-network (DQN), a reinforcement learning (RL) algorithm.

RL is an area of Artificial Intelligence (AI) where optimal policies are identified through a learning process that involves trial and error. This iterative process includes, at each step, an agent based on the current system *state* taking an *action* that causes the system to transition to another state that is associated with a certain *reward* [6, 7]. Q-learning algorithm is an RL algorithm that builds a Q-table to store, for every discretized *state(row)-action(column)* pair its estimated Q-values, which is a function of all future rewards. For every *state* the action that gives the maximum Q-value would then be the optimal action. Though it has vast successful applications, using a Q-table is feasible only for problems with discrete state space or continuous state spaces that can be discretized into a finite number of states [8]. For large state and action space environments where discretization is not sufficient or not feasible, an artificial neural network can be trained to learn the Q-values as a function of a continuous state space [8].

Deep Q-network (DQN), is such an algorithm where Q-tables are replaced with deep artificial neural networks (neural network with more than three layers, including input and output). In DQN, input nodes of the neural network are the current *state*, and the output nodes are the Q-value for each *action*. DQN was initially introduced in [9] and has been applied to a diverse set of problems, including but not limited to games [10], autonomous driving [11], recommendation system [12], mobile robot navigation [13], computer-aided diagnosis [14], stock trading [15], and very recently on COVID-19 pandemic control [15, 16].

In recent times, as lockdowns had been a key tool to curb COVID-19 spread, a growing number of RL studies have focused on identifying optimal lockdown policies with objective to minimize COVID-19 cases while also minimizing economic damages. In a study by Khadilkar et al., RL is used to automate policy learning to optimize lockdown policies for epidemic control [18]. Kompella et al. [19] developed an agent-based pandemic simulator and an RL-based methodology to optimize fine-grained mitigation policies that minimize the economic impact without overwhelming hospital capacity. In a study by Arango et al., RL is used to optimize cyclic lockdowns as a temporary alternative to extended lockdowns using two concurrent goals of minimizing overshoots of ICU usage and a socio-economic goal that minimizes the time spent under lockdown [20].

We present an RL model trained using the DQN algorithm to evaluate the question of whether a lockdown is necessary, and if so, when it should be initiated, to what level (proportion lockdown), and how it should change over time, such that it minimizes both epidemic and economic burdens. Though this objective is similar to other RL studies in the literature, our work differs from previous work in two ways. First, we evaluated the question of when to initiate a lockdown policy, which would be helpful for future outbreaks of similar epidemiology when lockdowns are a key intervention. Second, we evaluate these decisions in the context of two-geographical jurisdictions that make autonomous, independent decisions, cooperatively or non-cooperatively, but populations interact in the same environment through travel. Though decisions are made independently, because of travel between jurisdictions, the actions of one jurisdiction can influence the epidemic in the other jurisdiction. This scenario would especially be of interest for a jurisdiction that makes the optimal decisions but has travels coming from a jurisdiction with bad decisions. While travel between jurisdictions would be favorable for the economy, it could diminish the impact of its optimal actions. Therefore, taking the perspective of a jurisdiction that makes the optimal decision, we evaluate under travel when actions of another jurisdiction significantly add to its disease and economic burdens. This would help inform when border closures would need to be part of an optimal lockdown strategy. And subsequently, whether decision-making control should be given to individual jurisdictions (say county-level or state-level) or a common entity (such as state if jurisdictions are counties, and federal if jurisdictions are states). In this study, we assume that both jurisdictions start an outbreak at the same time, thus our results are limited to this scope.

The rest of the paper is organized as follows. Section 2 presents the methodology, including the simulation model, MDP formulation, and RL. In section 3, we discuss the scenarios we analyzed in detail. Section 4 presents the results, and finally, in section 5, we conclude the study with a discussion.

## 2. Methodology

Our model framework includes a compartmental simulation model that simulates the epidemic spread discussed in section 2.1 integrated with a Markov decision process (MDP) optimization framework discussed in section 2.2 and solved using deep Q-network (DQN) discussed in section 2.3.

### 2.1 Simulation Model

We developed a susceptible(S)-exposed(E)-infected(I)-recovered(R)-dead(D) (SEIRD) compartmental model based on Kermack and McKendrick [21] for simulating epidemic projections over time (Figure 1). An individual starts in compartment *S*, and upon contracting the disease moves to compartment *E*. A person in compartment *E* is in the incubation phase of the disease (for a duration of 1/*α* days) and thus cannot transmit the disease. A person moves from compartment E to compartment I, the transmissible phase of the infection. A person in compartment *I* either recovers, i.e., moves to *R* with rate *γ* per day, or succumbs to disease, i.e., moves to *D* with rate *θ* per day.

**Figure 1.**
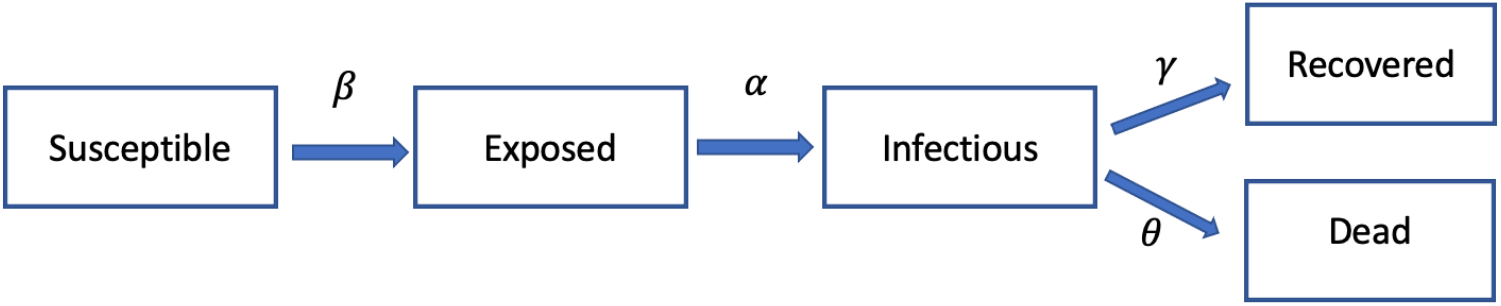
SEIRD flow diagram for infectious diseases.

Let

*S* be the number of Susceptible,

*E* be the number of Exposed,

*I* be the number of Infectious,

*R* be the number of Recovered,

*D* be the number of Dead,

*N* be total population,

*β*: transmission rate from susceptible to infected (*β* = *pc* where *p* is the probability of transmission per susceptible-infected contact and *c* =number of contacts per person),

*α*: is the inverse of the average incubation period in days,

*γ*: rate of recovery per day, and

*θ*: rate of disease-related mortality per day.

Given the short duration of the disease, we evaluate over a short analytic period of 400 days, assuming no births or natural deaths, and thus, the population size remains constant over time (*N* = *S*(*t*) + *E*(*t*) + *I*(*t*) + *R*(*t*) + *D*(*t*)). The differential equation governing the dynamics of the disease can be written as follows:

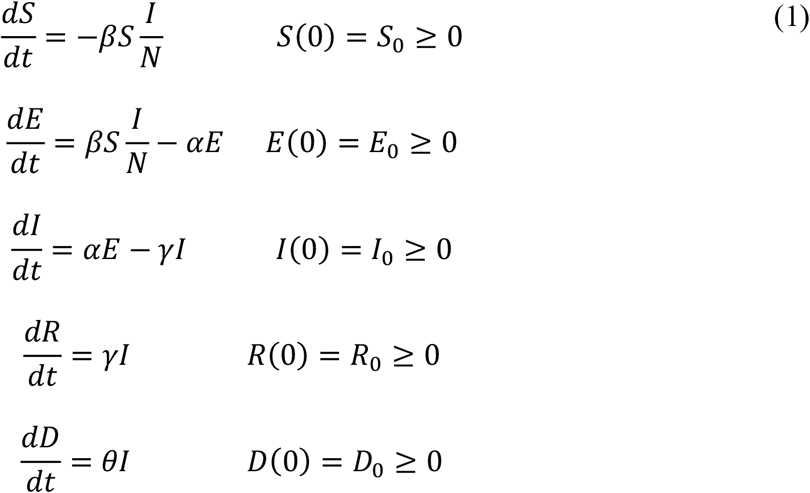

#### Population Mixing

To study the impact of travel on epidemic projections, we modified the standard SEIRD equations to include travel between two jurisdictions (jurisdiction A and jurisdiction B).

Let

*r*_*AB*_ be the travel rate from jurisdiction A to jurisdiction B,

*r*_*BA*_ be the travel rate from jurisdiction B to jurisdiction A, and

*I*_*B*_ be the number of infectious people in jurisdiction B.

Then the SEIRD model can be modified to include population mixing as follows:

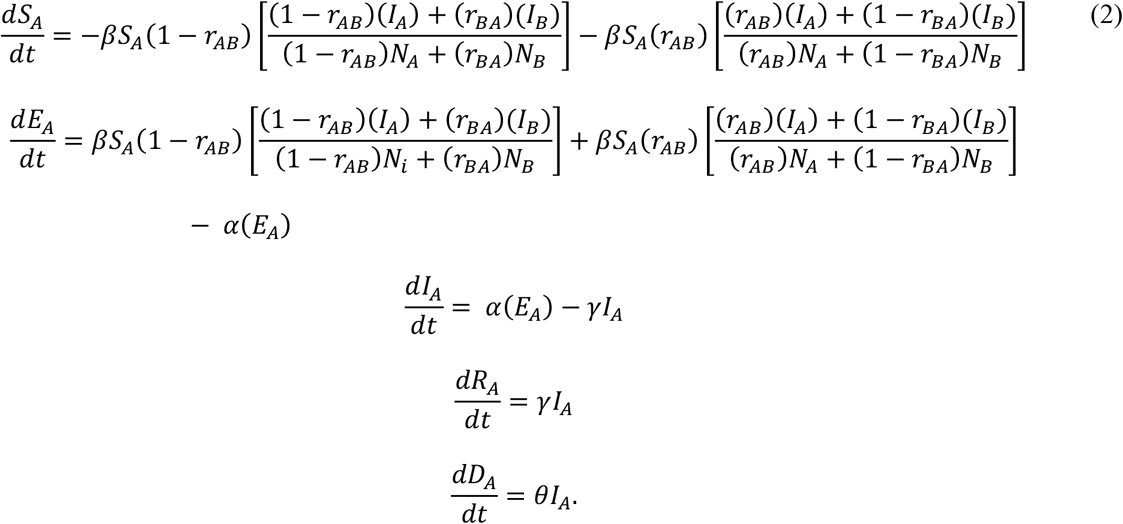

Note that setting *r*_*AB*_ = *r*_*BA*_ = 0 in (2) results in (1), and hence the single jurisdiction model is a special case of the two-jurisdiction model. For empirical analyses, we used epidemiology data from the SARS-Cov2 alpha variant (Table 1).

**Table 1.**
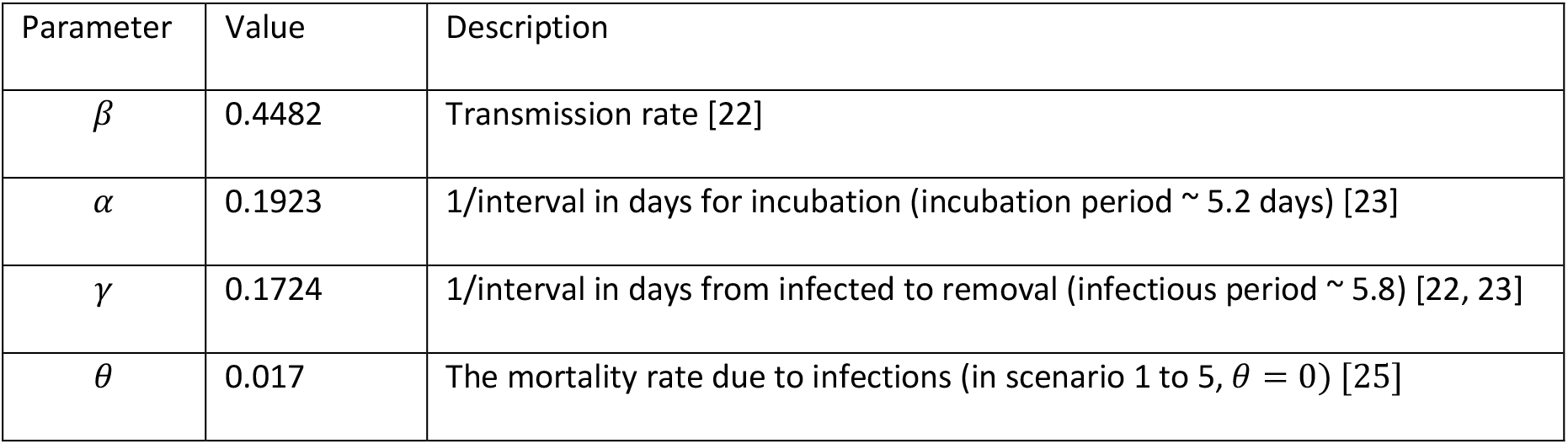
Parameters of the simulation model.

### 2.2 Markov Decision Process

We formulate the question of whether a lockdown is necessary, and if so, when it should be initiated, to what level (proportion lockdown), and how this should change over time as an MDP, as follows. We define the pandemic state as a multivariate parameter 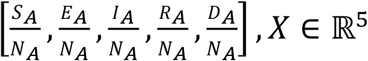, where 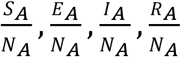, and 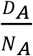 are the proportion of the jurisdiction A population in the S, E, I, R, and D compartment, respectively, and add to 1.

Then, using the standard form, we can define the MDP as a 5-tuple {Ω, 𝒜, *P*_*a*_, *R*_*a*_, *γ*}, where,

- Ω is the state space, a set of all possible states of the pandemic, *X ∈* Ω,
- 𝒜 is the action space, a set of all possible actions, here choices of lockdown, *a ∈* 𝒜,
- *P*_*a*_ is the one-step transition probability matrix from one state of pandemic to another under action *a* (where *P*_*a*_(*x*′|*a, x*) is the transition probability from state *x* to *x*′ under action *a*),
- ℛ_*a*_is a reward matrix, with each element, ℛ_*a*_(*x*′|*a, x*), the immediate reward of transitioning from state *x* to *x*′ under action *a*, and
- *γ* is the discount factor.

Given the system is in state *x*_0_ ∈ Ω at time of implementation of decision, the problem is to solve for the optimal policy (***d***(*x*_0_)) using the following objective function to maximize the total expected reward over the analytic period *T* (for numerical analyses we assumed *T* = 400):

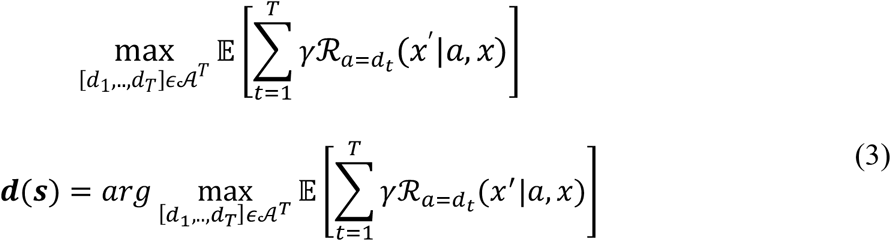

We next discuss the formulation of the 5-tuple {Ω, 𝒜, *P*_*a*_, ℛ_*a*_, *γ*}:

#### State space

We formulate the state space as 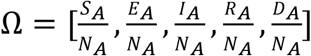, a continuous state space where each element of the state space can get a value between 0 and 1, such that at each time step, 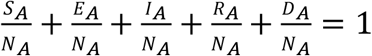.

#### Action space

We formulated the action space (𝒜) as a finite discrete set of interventions, 𝒜 = [*a*_1_ = 75%, *a*_2_ = 50%, *a*_3_ = 25%, *a*_4_ = 0%], corresponding to a contact rate reduction of 75%, 50%, 25%, and 0%, respectively, a factor multiplied to the transmission rate (*β*) in (1) and (2). For these numerical analyses, to make it representative of the COVID-19 epidemic, we assumed contact reductions are achieved through lockdowns. We assumed about 25% of the U.S. population are essential personnel [25, 26] (34% of adults reported as essential personnel, and 78% of the population are adults) and thus the strictest lockdown, *a*_1_, corresponds to a 75% reduction in contact rate. Value of action *a*_4_ was selected to represent no-lockdowns, and values of actions *a*_2_ and *a*_3_ were set at intermediate levels between *a*_1_ and *a*_4_.

#### Transition probabilities

As generating the transition probability for every possible transition is infeasible, we use our SEIRD simulation model discussed earlier to simulate each action and keep track of each transition in the model.

#### Immediate rewards

Immediate reward (ℛ_*a*_(*x*)) corresponds to the per time step reward (benefits – costs) achieved by implementing an *action* when the system is in *state x*. We evaluated immediate reward ℛ_*a*_(*x*) under two objective functions:

- 2-term objective function: The objective is to minimize economic burden and hospital capacity violation. This objective function would be most suitable for diseases that have a high risk of hospitalization, but minimal risk of mortality.
- 3-term objective function: The objective is to minimize economic burden, hospital capacity violation, and minimize mortalities. This objective function would be most suitable for diseases with high risk of hospitalizations and mortality.

Mathematically, we formulated the immediate reward ℛ_*a*_(*x*):

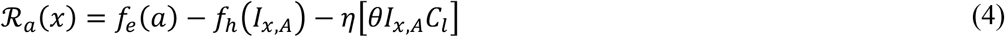

where, setting 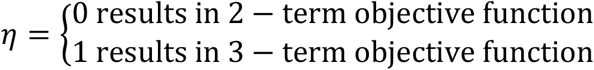,

*f*_2_(*a*) is the per day monetary benefit of implementing action *a*,

*f*_*h*_(*I*_*x,A*_) is the per day cost of exceeding hospital capacity in jurisdiction *A*, when there are *I*_4,*A*_ number of infected persons,

*θ* is the mortality rate, and thus *θI*_4,*A*_ is the number of daily deaths in jurisdiction *A* when there are

*I*_*x,A*_ number of infected persons, and

*C*_*l*_ is the per person mortality cost.

We modeled the monetary benefit (*f*_*e*_(*a*)) as the economic benefit,

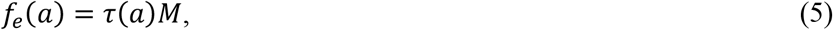

where, τ(*a*) is the monetary reduction in the economy upon implementation of action *a* and *M* is the per day monetary value generated by the economy in a no-lockdown scenario. Here, we assumed *M* = 1*e* + 11, and set τ(*a*_1_) = 0.4, τ(*a*_2_) = 0.6, τ(*a*_3_) = 0.8, and τ(*a*_4_) = 1. Per day monetary value of *M* is assumed based on US gross domestic product (GDP) per capita multiplied by US population in 2020 [28].

We assumed that for every 1000 inhabitants, there is 1.5 hospital beds available (we used data in the state of Utah which has the lowest number of beds per capita among US sates [28]) 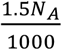 and that 5% of infected people at each timestep are hospitalized [21, 24], and modeled the per day cost of exceeding hospital capacity *f*_*h*_(*I*_*x,A*_) as

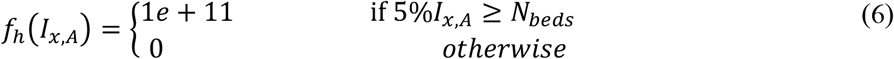

We assumed mortality rate is 0.017 corresponding to the SARS-Cov2 virus [25], and the cost per mortality (*C*_*l*_) as 1*e* + 10.

### 2.3 Deep Reinforcement Learning

We solve for the optimal sequence, level, and time of initiation of lockdowns for the control of COVID-19 type new infectious disease outbreaks, formulated above as an MDP, using DQN. We solve for this under varying scenarios (see Section 3). DQN is a deep reinforcement learning algorithm suitable for continuous state and discrete action spaces [9]. Conceptually, the algorithm works as follows. At each time step, based on the state of the pandemic, i.e., values for 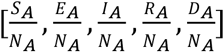, the algorithm determines what action to take, feeds it to the simulation model to calculate the immediate reward of taking that action at that particular state. This process is repeated for multiple iterations, and at every iteration, through training of a neural network, the algorithm is learning to take better actions, such that, under the proper neural network architecture and hyper-parameters, the algorithm eventually learns to identify the decision that maximizes the objective function defined in (3). We developed the model using the stable_baselines library in Python [28]. The details of the algorithm are presented in Appendix Section A.1.

#### DQN configuration and hyper-parameters

To approximate the Q-function, we used a deep learning network, a multi-layer perceptron with four layers that have 64, 128, 128, and 8 nodes, respectively. We use *γ*=0.95 and a learning rate of 0.001 with buffer size 100000. The rest of the parameters are set as default by the stable_baselines DQN library [28]. We trained each scenario separately for different number of MDP iterations (referred to as episodes), each 100 times with different random seeds.

The initial state at the beginning of each episode is set to one person exposed for jurisdiction *A* and two persons exposed for jurisdiction *B*, and rest of the population are susceptible. Each episode is 400 days, and at the end of each episode, the model is reset to the initial state. We trained the model for different episodes from 2500 to 25000 (corresponding to 1M to 10M time-steps). At the end of the training, we identify the optimal solution as the best among all the trained models, i.e., the model with the highest expected total reward (defined in (3)).

Similar to many optimization problems, DQN does not guarantee reaching the optimal solution, however, by sufficiently exploring the solution space, the chance of finding an optimal solution could be increased. Therefore, for each scenario (Section 3), we generated 100 different runs of the algorithm, each with a different random seed, and identifying an optimal solution under each. Similar optimal solutions in multiple runs would also suggest higher chance of optimality.

## 3. Analyses Scenarios

We analyzed seven scenarios. Scenario 1 to 5 correspond to the 2-term objective (that considers impact of decisions on economy and hospital capacity violation), while scenarios 6 and 7 correspond to the 3-term objective (that consider the impact of decisions on economy, hospital capacity violation, and disease related mortality). Scenarios 1 and 6 correspond to a single jurisdiction while the rest of the scenarios correspond to two-jurisdictions with different travel rates. In the two-jurisdiction scenarios, decisions are made independently, and we consider two distinct behaviors among them. In scenarios 2 and 3, jurisdiction A implements the optimal policy but jurisdiction B does not implement any intervention (non-cooperative behavior), while in scenario 4 and 5, jurisdiction B follows the exact same policy as A (cooperative behavior). However, note that, even in Scenarios 4 and 5, just as in Scenario 1 to 3, the formulation of the DQN focused only on the epidemic state in jurisdiction A. Thus, the DQN here was still a single-agent RL but evaluated in the context of two interacting jurisdictions making autonomous independent decisions. We further expanded these scenarios into sub-scenarios by examining the impact of delay in initiation of optimal policy, i.e., delaying initiation of optimal policy until day 30, 45, 60, 75, 90, 95, 100, 105, and 110 such that each corresponds to different prevalence upon initiation of optimal policy.

Intuitively, if the optimal policy is a lock-down, the more the delay in initiation of lockdown, the more the epidemic burden, but less of an economic burden. On the other hand, if the optimal policy is no-lockdown, it is equivalent to doing nothing, and so a delay in implementing optimal policy would not have any consequences until it reaches a time where the optimal policy shifts to a lockdown. Thus, the model technically considers the impact of delay and the tradeoff between economy and epidemic burden into its evaluation. Hence, the resulting optimal policy would also hold the answer to when a shutdown should be initiated. Besides, in the case of open borders, the optimal policy also changes based on the epidemic in the jurisdictions that the population interacts with through travel. However, the results would depend on how much weight (costs) is given to each objective function component. These costs associated with hospital capacity and lockdowns are likely to be subjective. For example, a jurisdiction where a significant fraction of jobs can seamlessly transition to remote work (e.g., IT) may differently weigh each of the four lockdown options (e.g., fewer days but maximum lockdown-level) compared to a jurisdiction where a large fraction of the jobs require physical presence (e.g., manufacturing) (e.g., extend days of lockdown at low lockdown-levels on each day). On the other hand, an infectious disease that is not deadly may be weighed lower for disease burden (hospital capacity as proxy) than a more deadly disease. Therefore, we made ‘time to initiate’ the optimal policy as an exogenous variable and evaluated multiple values. Details of the scenarios are discussed in Table 2.

**Table 2.** Summary of the scenarios studied.

| Scenario | Objective function | Number of jurisdictions | Policy | Travel from B to A | Initiation of optimal policy (days) |
| --- | --- | --- | --- | --- | --- |
| Scenario 1 | 2-term | Single jurisdiction, A | A optimal policy | Not applicable | 30, 60, 75, 90, 95, 100, 105, 110 |
| Scenario 2 | 2-term | Two jurisdictions, A and B | A optimal policy,<br>B no intervention | 5% | 30, 60, ..., 110 |
| Scenario 3 | 2-term | Two jurisdictions | A optimal policy,<br>B no intervention | 10% | 30, 60, ..., 110 |
| Scenario 4 | 2-term | Two jurisdictions | A optimal policy,<br>B optimal policy | 5% | 30, 60, ..., 110 |
| Scenario 5 | 2-term | Two jurisdictions | A optimal policy,<br>B optimal policy | 10% | 30, 60, ..., 110 |
| Scenario 6 | 3-term | Single jurisdiction | A optimal policy | Not applicable | 30, 60, ..., 110 |
| Scenario 7 | 3-term | Two jurisdictions | A optimal policy,<br>B no intervention | 10% | 30, 60, ..., 110 |

For each scenario, 1 to 7, we present the following metrics: the frequency of occurrence of each action over a 400-day period, the total number of days hospitalizations exceeded hospital capacity (which we will refer to as “hospital capacity violation”), number of hospitalizations, and additionally for Scenarios 6 and 7, the number of deaths.

We present the ‘initiation of optimal policy’ in days, which is how it was modeled, but also present the corresponding disease states, specifically, the observed prevalence and the actual prevalence. We define observed prevalence as the cumulative number of reported cases, tracked as part of disease surveillance, and expressed as a percentage of the total population. We define actual prevalence as the cumulative number of infected cases, i.e., it additionally includes those cases that are not yet reported and expressed as a percentage of the total population. Therefore, while the ‘initiation of optimal policy’ was modeled in days, the corresponding observed prevalence is more relevant and trackable from a public health perspective. In the case of the SARS-CoV2 virus, persons in the ‘*exposed*’ compartment are asymptomatic, and only show symptoms when they transition to the ‘*infectious*’ compartment. Therefore, we made a simplifying assumption that the observed prevalence includes all cases except those in the *exposed* compartment (i.e., includes infectious + recovered + death compartments), while the actual prevalence also includes the *exposed* compartment.

Note that, while all scenarios were modeled with the same time-points for ‘delay in initiation’, the epidemic projections under the different travel rates would be different and thus the corresponding values of observed prevalence and actual prevalence would vary by scenarios. For instance, 90 days of delay in scenario 1 corresponds to an observed prevalence of 1.35% and the actual prevalence of 2.13%, while the same days of delay in scenario 3 correspond to an observed prevalence of 1.9% and an actual prevalence of 3%. Therefore, we represent each sub-scenario, as [delay in initiation (in days), observed prevalence, and actual prevalence].

## 4. Results

In all scenarios, as expected from the highly virulent SARS-CoV2 virus, the optimal scenarios involved some lockdown until a majority of the population became infected or lasted for the entire simulation duration. In the 2-objective function scenarios (Scenarios 1 to 5), the optimal lockdown strategy helped avoid hospital capacity violations while minimizing the economic burden from lockdowns by taking the least stringent lockdown. However, the optimal policy was to end lockdown only after a majority of the population became infected and reached herd-immunity levels. In the 3-objective function scenarios (Scenarios 6 and 7), the optimal lockdown strategy helped avoid hospital capacity violations, minimize infected cases and deaths while minimizing the economic burden from lockdowns by taking the least stringent lockdown. However, the optimal strategy here was to continue the optimal pattern of lockdowns for the remaining duration of the simulation, suggesting that until a vaccine becomes available, there is a chance that the infection would spread. We discuss these results in more detail below.

With only one jurisdiction (Scenario 1), the optimal strategy was to initiate lockdown if the observed prevalence (proportion of the population infected) reached 2.3% (which corresponded to the actual prevalence of 3.6%). This can be seen in Figure 2 (first row), scenarios where lockdown initiated at the observed prevalence of 2.3% or below (corresponding to up to 95 days from time of first case) had least lockdown and similar outcome of zero hospital capacity violations. Over the duration of 400 days, this optimal policy consisted of lockdown at 50% for 62 days and lockdown at 25% for 46 days. Under this policy, lockdowns could be fully lifted on day 209. In the optimal strategy, the number hospitalized per day never exceeded hospital capacity, i.e., zero days of hospital capacity violation. As expected from including only economy and hospital capacity in the objective function, given the high infectiousness of the virus and absence of other interventions, about 79% of the population were infected over the duration of the pandemic Figure 3 (first row).

**Figure 2:**
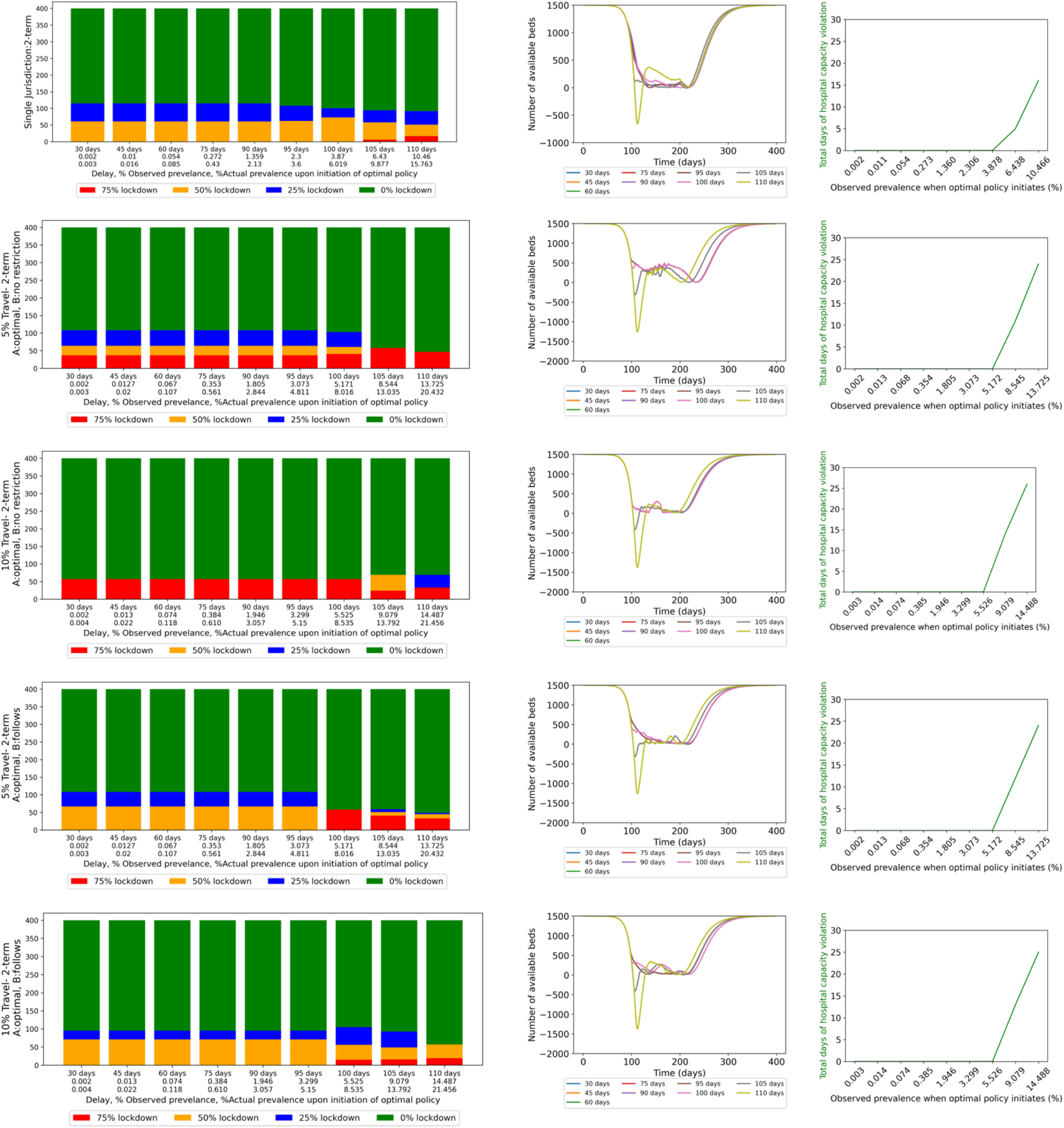
2-term objective function models for scenarios 1, 2, 3, 4, and 5. Left plots: Bar plots of frequency of occurrence of each action (75% (red), 50% (yellow), 25% (blue), and 0% (red) lockdown) over 400 days for different delay (x-axis) in initiation of optimal policy [delay in days, observed prevalence, and actual prevalence]. Middle plots: Number of available hospital beds (y-axis) against time (x-axis) under different delays in initiation of optimal policy. Right plots: Total number of days hospital capacity is violated (y-axis) against observed prevalence at time of initiation of optimal policy (x-axis).

**Figure 3:**
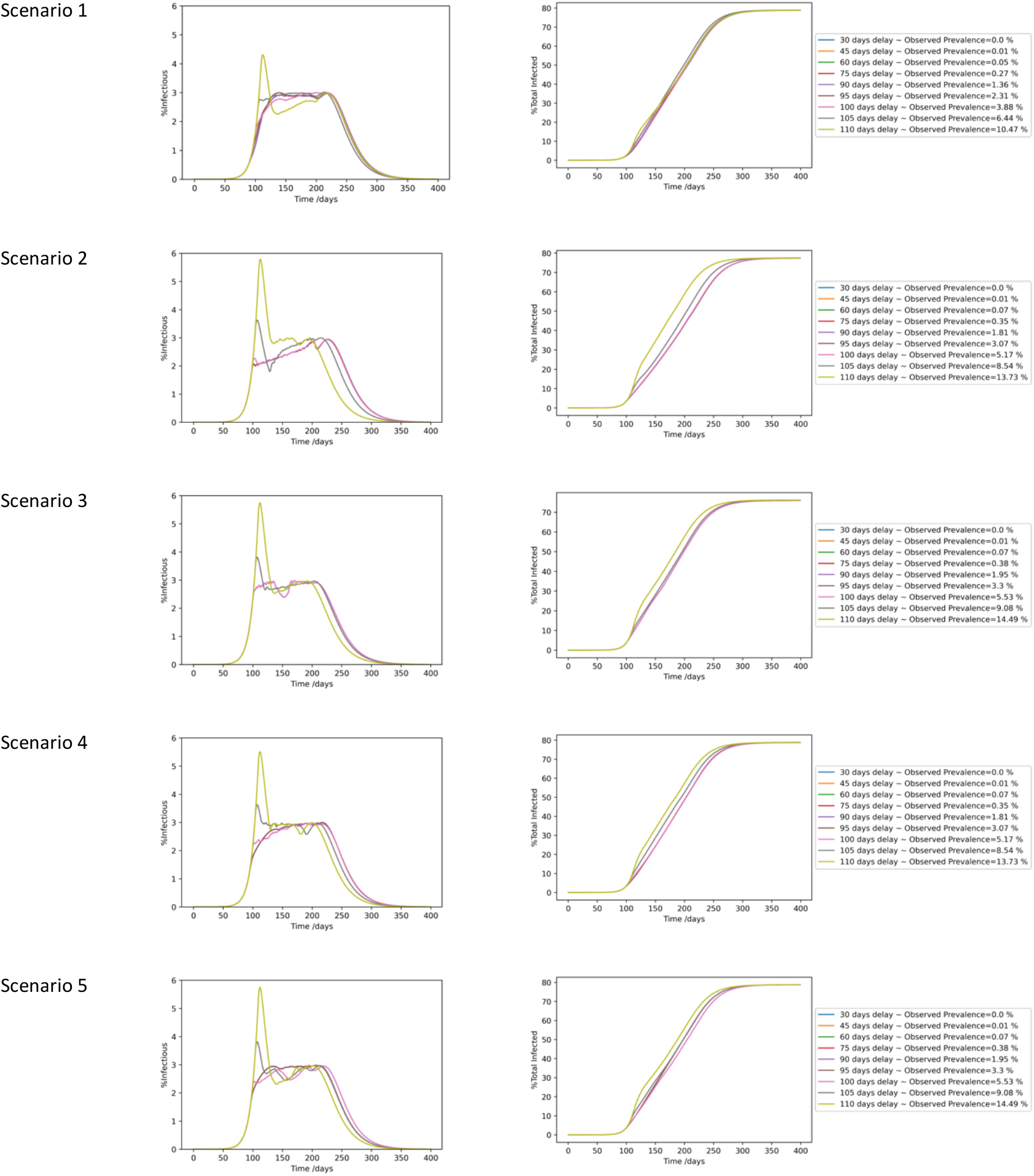
Percentage infectious among total population vs time for different delays in initiation of the optimal policy (left plots) and corresponding impact on percentage total infected over time (right plots) for scenarios 1, 2, 3, 4, and 5.

Delaying implementation of optimal policy in Scenario 1, i.e., initiating lockdown after observed prevalence exceeded 2.3%, led to more prolonged or more stringent lockdowns and/or hospital capacity violations (Figure 3 first row). For example, delaying to until 3.8% observed prevalence led to 73 days of 50% shutdown, 27 days of 25% shutdown, and zero days of hospital capacity violation. Delaying to 6.4% observed prevalence led to 6 days of 75% shutdown, 52 days of 50% shutdown, 36 days of 25% shutdown, and five days of hospital capacity violation. Delaying to until 10.46% observed prevalence led to 17 days of 75% shutdown, followed by 34 days of 50% shutdown, 41 days of 25% shutdown, and 16 days of hospital capacity violation. While the 1.35% observed prevalence occurred on day 90, the observed prevalence of 2.3%, 3.88%, 6.43%, and 10.46% occurred on days 95, 100, 105, and 110, suggesting that because of the high infectiousness of the virus, a few days of delay could lead to significantly worse disease and economic burdens.

When jurisdiction A interacted with jurisdiction B through travel, but jurisdiction B was non-cooperative and did not take the optimal decision as A (Scenarios 2 and 3 –with 5% and 10% travel, respectively), the optimal policy for A was to control for B’s non-cooperative actions through more stringent lockdowns than in Scenario 1 (0% travel). Even with the lower 5% travel (Scenario 2-Figure 2 second row) and initiating lockdowns when observed prevalence was as low as 0.002% (30 days delay), unlike in Scenario 1 (Figure 2 first row), the optimal lockdown involved 28 days of maximum 75% lockdown.

In Scenario 2, the optimal lockdown strategy up until observed prevalence of 3.07% were similar with outcomes of zero days of hospital capacity violation. The optimal policy, over the period of 400 days, was lockdowns at the maximum-level of 75% for 37 days before transitioning to the less stringent 50% and 25% levels. Delayed implementation of optimal policy until the observed prevalence reached 5.17% led to the need for more stringent lockdowns (41 days of the maximum 75%, 20 days of 50%, and 42 days of 25%) to avoid hospital capacity violation. Delaying implementation of optimal policy to beyond observed prevalence of 5.17% led to a situation where hospital capacity violations could not be avoided (Figure 2 second row). For example, delaying until 8.54% observed prevalence led to 58 days of 75% shutdown, and 11 days of hospital capacity violation. Delaying to until 13.73% observed prevalence led to 47 days of 75% shutdown, and 24 days of hospital capacity violation.

In Scenario 3 (Figure 2 third row), the optimal policy was to initiate a lockdown no later than an observed prevalence of 5.52%. The optimal policy, over the period of 400 days, was lockdowns at the maximum level of 75% for 57 days, which resulted in zero days of hospital capacity violation. Delaying implementation of optimal policy to after observed prevalence exceeded 5.52%, led to higher hospital capacity violations (Figure 2 third row). For example, delaying until observed prevalence was 9.07% led to 24 days of 75% shutdown, followed by 45 days of 50% shutdown, and 14 days of hospital capacity violation. Delaying until observed prevalence was 14.48% led to 33 days of 75% shutdown, followed by 36 days of 25% shutdown and 26 days of hospital capacity violation.

When jurisdiction A interacted with B through travel but unlike the above scenarios, B was cooperative by taking optimal actions as A (Scenarios 4 and 5), the optimal policy was similar to that in Scenario 1 (single jurisdiction, 0% travel), suggesting that cooperative behavior would yield similar results as single jurisdiction, as expected. Note that, similarity in results between Scenarios 4, 5, and 1 suggests that, though the DQN was trained as a single-agent RL by considering only the state space of jurisdiction A, this is a sufficient method here as we assumed that both jurisdictions start the epidemic at the same time.

In summary, results from the above 2-objective function scenarios suggest that deviating from the optimal policy through delays in initiating the optimal policy or through non-cooperative behavior by an outside but interacting jurisdiction (B in this case) would require more stringent lockdowns (red bar) to avoid hospital capacity violations.

With the 3-objective function, and only one jurisdiction (Scenario 6), the optimal policy was to initiate lockdown when observed prevalence was 0.01% (Figure 4 first row). Under this, the optimal lockdown policy continued for the remaining duration of the simulation in order to reduce cases and keep deaths at zero. This suggests that until pharmaceutical options are available, preventing highly transmissible diseases such as COVID-19 would require some level of physical distancing between contacts. Delaying the initiation of the optimal policy generated multiple deaths even though higher number of lockdowns were initiated to control for the delays. Delaying implementation of an optimal strategy to prevalence 10.46% (which occurred on day 110 from the first infection) resulted in 4374 deaths, and 16 hospital capacity violations (Figure 4 first row, and Appendix Table A2).

**Figure 4:**
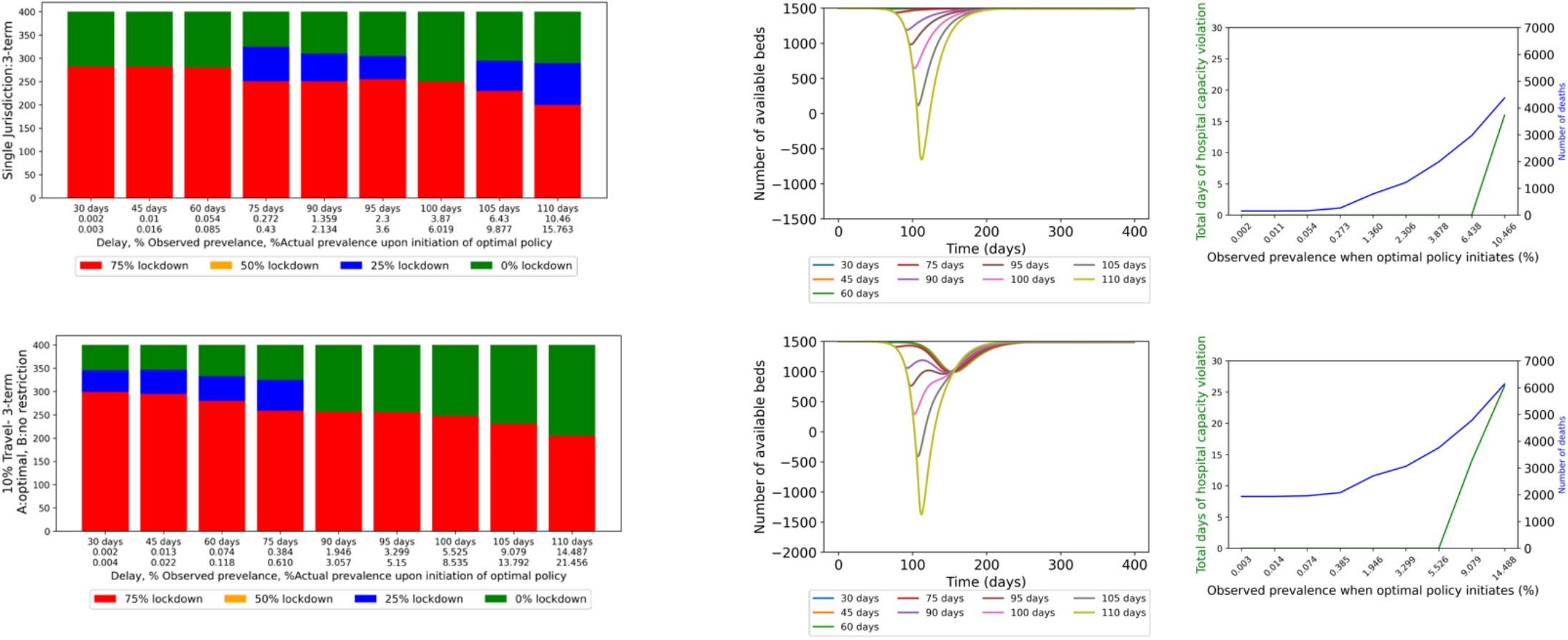
3-term objective function models for scenarios 6 and 7. Left plots: Bar plots of frequency of occurrences of each action (75% (red), 50% (yellow), 25% (blue), and 0% (red) lockdown) over 400 days for different delays (x-axis) in initiation of optimal policy]delays in days, observed prevalence, and actual prevalence]. Middle plots: Number of available hospital beds (y-axis) against time (x-axis) under different delays in initiation of optimal policy. Right plots: Total number of days hospital capacity is violated (y-axis) against observed prevalence at time of initiation of optimal policy (x-axis).

**Figure 5:**
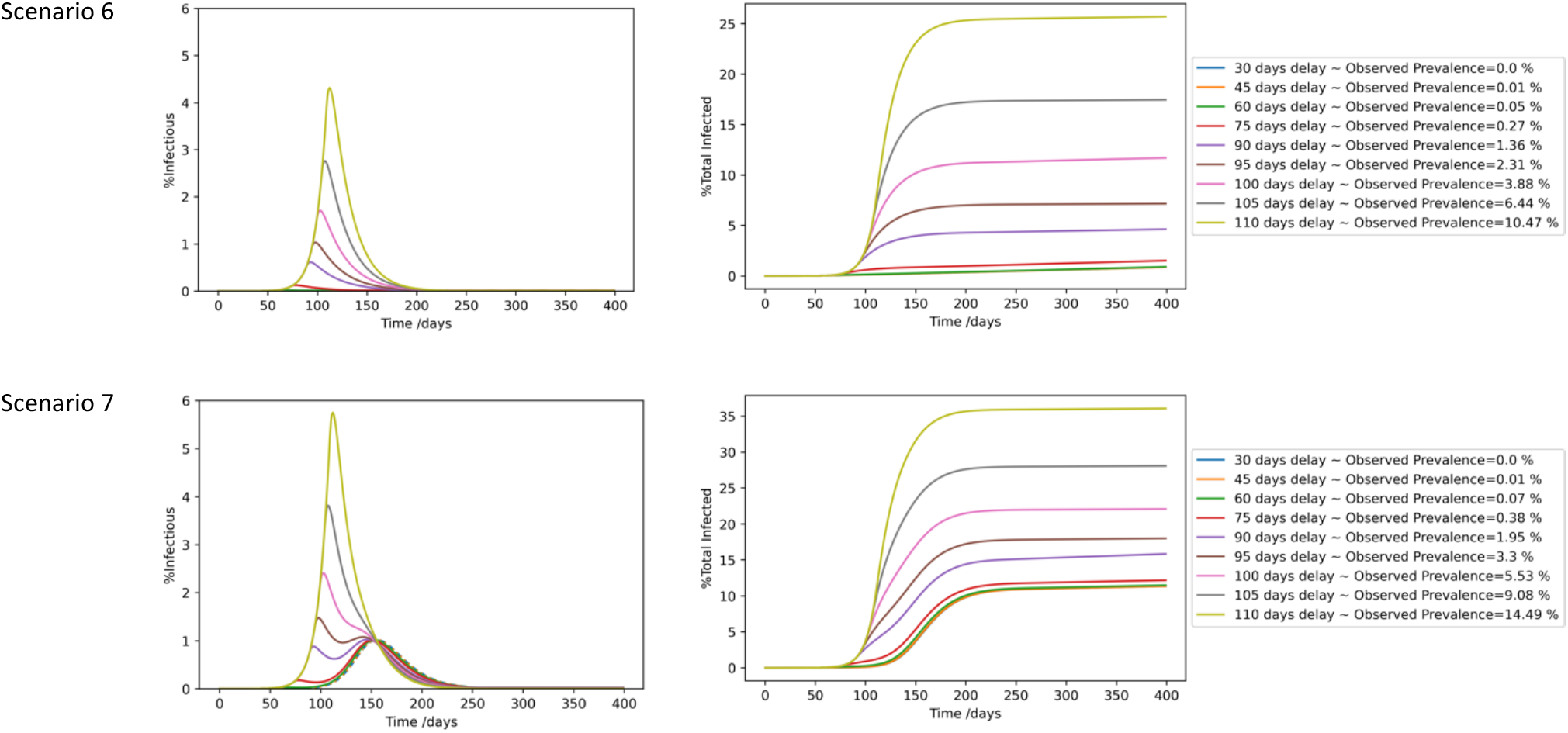
Percentage infectious among total population vs time for different delays in initiation of the optimal policy (left plots) and corresponding impact on percentage total infected over time (right plots) for scenarios 6 and 7.

With the 3-objective function, when jurisdiction A was interacting with B through travel, but jurisdiction B was not implementing any interventions (Scenario 7), the optimal strategy for A to control for the non-cooperative behavior of B were a greater number of days and more stringent lockdowns. Under this, the optimal policy over the 400 days was lockdown at the highest-level of 75% for 299 days and at 25% for an additional 47 days (Figure 4 row 2). This optimal policy resulted in zero days of hospital capacity violation but 1935 deaths (Figure 4 row 2). Delaying implementation of the optimal policy until observed prevalence reached 8.5%, led to a situation where the epidemic burden had already created sufficient deaths that lockdowns had a lesser impact and could only be implemented to reduce future deaths than to prevent deaths. The optimal policy in this case was 231 days of the highest-level of 75% lockdown and resulted in 4775 deaths and 14 days of hospital capacity violation.

### Comparing results between 2-objecive and 3-objectve functions

In the 2-term objective function, as the objective was to only minimize economic burden and hospital capacity violations, the cumulative prevalence reached up to 80%, (Figure 3) i.e., the main outcome was that it reduced daily cases sufficient enough to keep hospitalizations below hospital capacity. In the 3-objective function, as the objective additionally minimized deaths, even in the worst-case scenario the cumulative prevalence reached about 35%. However, a key consequence of this was that, while in the 2-objective function lockdowns could be lifted within the timeline of the simulation, in the 3-objective function lockdowns continued over the full duration of the simulation. This suggests the need for continuing shutdowns until the availability of pharmaceutical interventions such as treatment to prevent deaths or vaccines to prevent transmissions.

Details of optimal policy of each sub-scenarios of 2-objective and 3-objective are presented in Appendix Table A1.

## 5. Conclusion and Discussion

We formulated the question of how to control epidemics such as COVID-19 in the absence of pharmaceutical interventions as a sequential decision-making problem formulated as a Markov decision process (MDP) and solved using Deep Q-network (DQN), a reinforcement learning algorithm. We propose a methodology that can help determine whether and when a lockdown is necessary, to what level, and how to phase out a lockdown which is a critical part of a pandemic preparedness plan. Furthermore, we evaluated these decisions in the context of two-geographical jurisdictions that make autonomous, independent decisions, cooperatively or non-cooperatively, but interact in the same environment through travel. We evaluated these decisions both under a 2-term objective function that minimized economic burden and hospital capacity violations, suitable for diseases with high-risk of hospitalizations but low risk of mortality, and a 3-term objective function that additionally minimized deaths. We used a SEIRD model to simulate the disease progression and incorporated the impact of travel in the formulation of the transmission rate.

In the case of a single jurisdiction, under a 2-term objective, the optimal time for initiation of lockdowns would be at about an observed prevalence of 3.87% and included lockdowns at a combination of 50% and 25% per day. Delaying decisions led to a higher number or more stringent lockdowns at the maximum levels of 75% per day in addition to a higher number of hospital capacity violations. In the case of two-jurisdictions A and B interacting through travel, if jurisdiction B deviated from the optimal policy, jurisdiction A would have to implement more stringent lockdowns to compensate for the non-cooperative behavior of B, and if there was any delay in this implementation also face excess hospitalizations. This suggests that, even if jurisdictions make decisions independently, cooperation between jurisdictions could help minimize lockdowns and avoid border travel restrictions, thus minimizing overall economic burden. In the absence of such cooperation, the trade-offs for jurisdiction A to consider would be between more stringent lockdowns within its jurisdiction or border closures to remove the interactions with jurisdiction B. The results are intuitive, what the study contributes is a methodology that can be used by jurisdictions to evaluate a suitable policy, under such interactive environments, and the numerical analyses here serves as proof-of-concept for the method.

In the 2-objective function scenarios (Scenarios 1 to 5), the optimal lockdown strategy helped avoid hospital capacity violations while minimizing the economic burden from lockdowns by taking the least stringent lockdown. However, as expected from the high transmissibility of the virus, the optimal policy was to end lockdowns only after a majority of the population became infected and reached herd-immunity levels. In the 3-objective function scenarios (Scenarios 6 and 7), the optimal lockdown strategy helped avoid hospital capacity violations, minimized infected cases and deaths while minimizing the economic burden from lockdowns by taking the least stringent lockdown. However, the optimal strategy here was to continue the optimal pattern of lockdowns for the remaining duration of the simulation, suggesting that shutdowns would have to continue until a vaccine became available. Any deviations from this optimal policy generated more stringent lockdowns and/or higher cases of hospitalizations and deaths. This suggests that, in the absence of pharmaceutical interventions, some measures of physical distancing would be necessary to control the epidemic even if it creates economic burdens, as deviating from this would only increase future economic burdens.

Some of the limitations of our model are as follows. Motivated by the COVID-19 pandemic, for the numerical analyses, we assumed epidemiology staging and transmissibility of the SARS-CoV 2 virus. Thus, the specific results here are limited to diseases caused by viruses similar to that of SARS-CoV 2 type. The model will have to be reparametrized and evaluated for other diseases with vary epidemiology structures. In our model, the impact of lockdowns on the economy is scaled linearly, i.e., lockdown on any day has a similar impact on the economy’s monetary value. This impact can be formulated as a non-linear function to consider the dynamical changes over time. We assumed that both jurisdictions start an outbreak at the same time, thus it was sufficient to train the DQN as a single-jurisdiction RL with both jurisdictions implementing the same policy (as evident from the similarity in results between Scenario 4, 5, and 1). Thus, our results are limited to this scope. For evaluating decisions between two jurisdictions that start the outbreak at different times leading to significantly different states of the epidemic at the time of decision-making, other methods such as multi-agent RL maybe more relevant.

Despite these limitations, we believe that the methodology presented here can help decision makers in formulating a pandemic preparedness plan for future infectious disease outbreaks. The results generated by the numerical analyses are intuitive, which support the feasibility of application of AI algorithms for such analyses, as typically, given the computational complexity of the algorithms and problem formulation, the feasibility is not always guaranteed [29]. This study provides a generalized framework that can be applied to any jurisdiction or infectious disease by adjusting the parameters accordingly, some examples are as follows. We interpreted the intervention options here to represent lockdowns and did not consider other options such as facemask use, self-isolation when infected, or 6 ft distancing. However, we modeled lockdowns by reducing transmission rate, assuming that the cost for that reduction represents economic loss. Interventions such as facemask use, self-isolation when infected, or 6 ft distancing are also modeled as reduction in transmission rates, but they may differ in governmental lockdowns in terms of the cost and impact, i.e., they may have a lesser impact on the economy (lower costs) but also achieve a smaller reduction in transmission rate. Therefore, the different levels of shutdowns and costs modeled here can also be interpreted as different types of interventions and the corresponding transmission rate, rewards, and costs informed specific to the setting. Design of the reward function is an essential step in RL models and can significantly change the optimal policy. Thus, this is a subjective metric that should be informed specific to the case under study. For example, a jurisdiction where a significant fraction of jobs can seamlessly transition to remote work (e.g., IT) may differently weigh each of the four lockdown options (e.g., fewer days but maximum lockdown-level) compared to a jurisdiction where a large fraction of the jobs require physical presence (e.g., manufacturing, or essential workers). On the other hand, those costs saved from preventing economic loss could instead be redirected to ensure safety of workers. Thus, the reward function would be formulated to consider economic costs, epidemic costs, and costs for safety measures.

## Data Availability

All data produced in the present work are contained in the manuscript

## Abbreviations

NPI: Non-pharmaceutical Interventions
MDP: Markov Decision Process
DQN: Deep Q-Network
RL: Reinforcement Learning
SEIRD: susceptible(S)-exposed(E)-infected(I)-recovered(R)-dead(D)
GDP: Gross Domestic Product

## Acknowledgments

This material is based upon work supported by the National Science Foundation NSF 1915481. Any opinions, findings, and conclusions or recommendations expressed in this material are those of the author(s) and do not necessarily reflect the views of the National Science Foundation.

## APPENDIX

### A.1 Details of DQN Algorithm

Throughout the training, a Q-table gets updated in the Q-learning algorithm, where each table element represents a state-action value. In the case of ample state space, when building a Q-table is intractable, or in the case of continuous state space, a Q-function is used to map state-action pair to a Q-value. Deep neural networks are used as a function approximator of state-action pair to a Q-value. During training, the algorithm learns the weights of the deep neural network. As a result, given the state to the deep Q-network (DQN), the Q-value associated with each action is outputted. And hence, the highest Q-value corresponds to the optimal action.

In the case of this study, the trained Q-network works as follow:

**Figure F:**
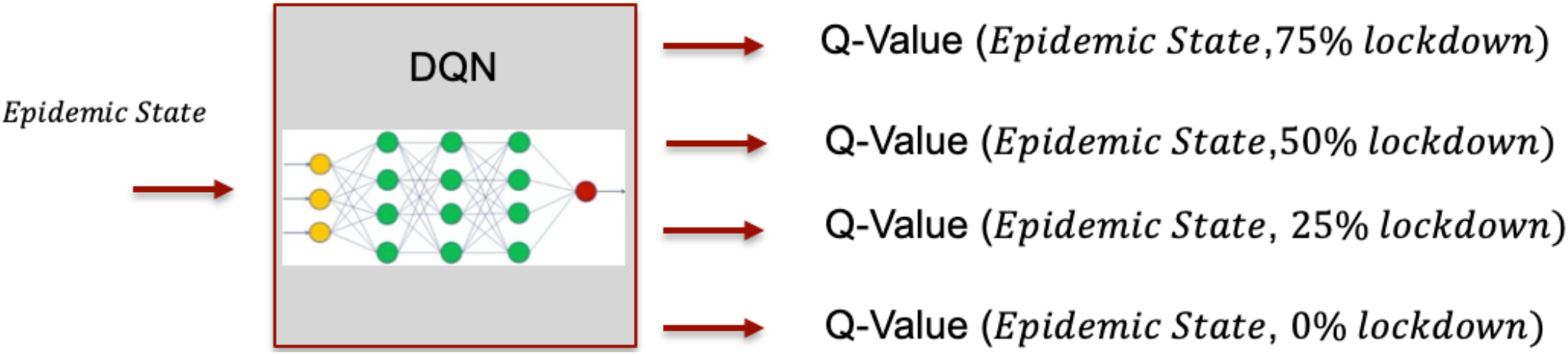
Schematic of DQN in the context of this study.

Given the epidemic state, the Deep Q-network outputs the Q-value associate to every action. And hence, the highest Q-value corresponds to the optimal action in that epidemic state.

In the DQN algorithm, there are two networks (two artificial neural networks), and an experience replay:

▪ Q-Neural network: is usually a deep neural network,
▪ Target Neural Network: is identical to the Q-Neural network,
▪ Experience replay: is used to memorize the agent’s experience when interacting with the environment, i.e. (state, action, next state, reward) as to reduce the correlation between the agent’s experiences and prevent overfitting of the network.

DQN algorithm is described as bellow [1]:

- Initialize replay memory to some capacity.
- Initialize Q-network with random weights.
- For a pre-defined number of episodes:
  - At each training step, random samples from experience replay are selected.
  - Batch of training data is fed into the Q-network and target network.
  - An action is taken based on the pre-defined action selection strategy, i.e., epsilon greedy.
  - DQN and target network separately predict the Q-values of current state and all the actions.
  - The experience is stored as a pair of (state, action, next state, reward) in the replay buffer.
  - Mean squared loss gets calculated based on the Q-value of target network and Q-network.
  - The loss gets backpropagated to the Q-network so to update the weights using gradient descent algorithm.
  - After a pre-defined number of time-steps, Q-network weights get copied to the target network. Q-network and target network become identical again.

### A.2 Summary of Optimal Policy for Scenarios 1 to 5

**Table A3:** Summary of scenarios 1 to 5. Shaded cells are the optimal policy in terms of when to be implemented (number of days of delays) and how to be implemented (number of days in each lockdown categories).

| Delays |  |  | 30 days | 45 days | 60 days | 75 days | 90 days |
| --- | --- | --- | --- | --- | --- | --- | --- |
| Scenario 1 | Observed Prevalence (%) |  | 0.002 | 0.01 | 0.054 | 0.272 | 1.359 |
|  | Actual Prevalence (%) |  | 0.0032 | 0.016 | 0.085 | 0.43 | 2.134 |
|  | Number shutdown | 0% | 285 | 285 | 285 | 285 | 285 |
|  |  | 25% | 54 | 54 | 54 | 54 | 54 |
|  |  | 50% | 61 | 61 | 61 | 61 | 61 |
|  |  | 75% | 0 | 0 | 0 | 0 | 0 |
|  | Number of days of hospital capacity exceeded |  | 0 | 0 | 0 | 0 | 0 |
| Scenario 2 & 4 | Observed Prevalence (%) |  | 0.0023 | 0.0127 | 0.0676 | 0.353 | 1.805 |
|  | Actual Prevalence (%) |  | 0.0037 | 0.02 | 0.1079 | 0.561 | 2.844 |
| Scenario 2 | Number shutdown | 0% | 292 | 292 | 292 | 292 | 292 |
|  |  | 25% | 44 | 44 | 44 | 44 | 44 |
|  |  | 50% | 27 | 27 | 27 | 27 | 27 |
|  |  | 75% | 37 | 37 | 37 | 37 | 37 |
|  | Number of days of hospital capacity exceeded |  | 0 | 0 | 0 | 0 | 0 |
| Scenario 4 | Number shutdown | 0% | 292 | 292 | 292 | 292 | 292 |
|  |  | 25% | 41 | 41 | 41 | 41 | 41 |
|  |  | 50% | 67 | 67 | 67 | 67 | 67 |
|  |  | 75% | 0 | 0 | 0 | 0 | 0 |
|  | Number of days of hospital capacity exceeded |  | 0 | 0 | 0 | 0 | 0 |
| **Scenario 3 & 5** | Observed Prevalence (%) |  | 0.0025 | 0.0139 | 0.0741 | 0.384 | 1.946 |
|  | Actual Prevalence (%) |  | 0.004 | 0.0223 | 0.118 | 0.610 | 3.057 |
| **Scenario 3** | Number shutdown | 0% | 343 | 343 | 343 | 343 | 343 |
|  |  | 25% | 0 | 0 | 0 | 0 | 0 |
|  |  | 50% | 0 | 0 | 0 | 0 | 0 |
|  |  | 75% | 57 | 57 | 57 | 57 | 57 |
|  | Number of days of hospital capacity exceeded |  | 0 | 0 | 0 | 0 | 0 |
| **Scenario 5** | Number shutdown | 0% | 305 | 305 | 305 | 44 | 305 |
|  |  | 25% | 24 | 24 | 24 | 27 | 24 |
|  |  | 50% | 71 | 71 | 71 | 37 | 71 |
|  |  | 75% | 0 | 0 | 0 | 0 | 0 |
|  | Number of days of hospital capacity exceeded |  | 0 | 0 | 0 | 0 | 0 |
| **Delays** |  |  | **95 days** | **100 days** | **105 days** | **110 days** | **120 days** |
| **Scenario 1** | Observed Prevalence (%) |  | 2.305 | 3.878 | 6.438 | 10.465 | 24.787 |
|  | Actual Prevalence (%) |  | 3.60 | 6.019 | 9.877 | 15.763 | 34.90 |
|  | Number shutdown | 0% | 292 | 300 | 306 | 308 | 368 |
|  |  | 25% | 46 | 27 | 36 | 41 | 1 |
|  |  | 50% | 62 | 73 | 52 | 34 | 11 |
|  |  | 75% | 0 | 0 | 6 | 17 | 20 |
|  | Number of days of hospital capacity exceeded |  | 0 | 0 | 5 | 16 | 36 |
| **Scenario 2 & 4** | Observed Prevalence (%) |  | 3.073 | 5.1716 | 8.544 | 13.725 | 30.86 |
|  | Actual Prevalence (%) |  | 4.811 | 8.016 | 13.035 | 20.432 | 42.29 |
| **Scenario 2** | Number shutdown | 0% | 292 | 297 | 342 | 353 | 353 |
|  |  | 25% | 44 | 42 | 0 | 0 | 0 |
|  |  | 50% | 27 | 20 | 0 | 0 | 0 |
|  |  | 75% | 37 | 41 | 58 | 47 | 47 |
|  | Number of days of hospital capacity exceeded |  | 0 | 0 | 11 | 24 | 58 |
| **Scenario 4** | Number shutdown | 0% | 292 | 342 | 341 | 351 | 373 |
|  |  | 25% | 41 | 0 | 8 | 4 | 6 |
|  |  | 50% | 67 | 0 | 10 | 12 | 1 |
|  |  | 75% | 0 | 58 | 41 | 33 | 20 |
|  | Number of days of hospital capacity exceeded |  | 0 | 0 | 12 | 24 | 41 |
| **Scenario 3 & 5** | Observed Prevalence (%) |  | 3.299 | 5.525 | 9.079 | 14.487 | 32.049 |
|  | Actual Prevalence (%) |  | 5.150 | 8.535 | 13.792 | 21.456 | 43.618 |
| **Scenario 3** | Number shutdown | 0% | 343 | 343 | 331 | 331 | 375 |
|  |  | 25% | 0 | 0 | 0 | 36 | 0 |
|  |  | 50% | 0 | 0 | 45 | 0 | 0 |
|  |  | 75% | 57 | 57 | 24 | 33 | 25 |
|  | Number of days of hospital capacity exceeded |  | 0 | 0 | 14 | 26 | 41 |
| **Scenario 5** | Number shutdown | 0% | 305 | 295 | 307 | 343 | 374 |
|  |  | 25% | 24 | 49 | 44 | 0 | 6 |
|  |  | 50% | 71 | 41 | 33 | 38 | 0 |
|  |  | 75% | 0 | 15 | 16 | 19 | 20 |
|  | Number of days of hospital capacity exceeded |  | 0 | 0 | 13 | 25 | 40 |

### A.3 Summary of Optimal Policy for Scenarios 6 and 7

**Table A4:** Summary of scenarios 6 and 7. Shaded cells are the optimal policy in terms of when to be implemented (number of days of delays) and how to be implemented (number of days in each lockdown categories)

| Delay |  |  | 30 days | 45 days | 60 days | 75 days | 90 days |
| --- | --- | --- | --- | --- | --- | --- | --- |
| Scenario 6 | Observed Prevalence (%) |  | 0.002 | 0.01 | 0.054 | 0.272 | 1.359 |
|  | Actual Prevalence (%) |  | 0.0032 | 0.016 | 0.085 | 0.43 | 2.134 |
|  | Number shutdown: | 0% | 118 | 118 | 120 | 75 | 90 |
|  |  | 25% | 0 | 0 | 0 | 74 | 59 |
|  |  | 50% | 0 | 0 | 0 | 0 | 0 |
|  |  | 75% | 282 | 282 | 280 | 251 | 251 |
|  | Number of days of hospital capacity exceeded |  | 0 | 0 | 0 | 0 | 0 |
|  | Number of deaths |  | 152 | 152 | 159 | 264 | 789 |
| Scenario 7 | Observed Prevalence (%) |  | 0.0025 | 0.0139 | 0.0741 | 0.384 | 1.946 |
|  | Actual Prevalence (%) |  | 0.004 | 0.0223 | 0.118 | 0.610 | 3.057 |
|  | Number shutdown: | 0% | 54 | 53 | 67 | 75 | 143 |
|  |  | 25% | 47 | 52 | 53 | 66 | 0 |
|  |  | 50% | 0 | 0 | 0 | 0 | 0 |
|  |  | 75% | 299 | 295 | 280 | 259 | 257 |
|  | Number of days of hospital capacity exceeded |  | 0 | 0 | 0 | 0 | 0 |
|  | Number of deaths |  | 1935 | 1936 | 1959 | 2077 | 2703 |
| **Delays** |  |  | **95 days** | **100 days** | **105 days** | **110 days** | **120 days** |
| **Scenario 6** | Observed (%) |  | 2.305 | 3.878 | 6.438 | 10.465 | 24.787 |
|  | Actual (%) |  | 3.60 | 6.019 | 9.877 | 15.763 | 34.90 |
|  | Number shutdown | 0% | 95 | 150 | 105 | 110 | 172 |
|  |  | 25% | 50 | 0 | 65 | 90 | 0 |
|  |  | 50% | 0 | 0 | 0 | 0 | 0 |
|  |  | 75% | 255 | 250 | 230 | 200 | 228 |
|  | Number of days of hospital capacity exceeded |  | 0 | 0 | 0 | 16 | 35 |
|  | Number of deaths |  | 1219 | 1992 | 2971 | 4374 | 7912 |
| **Scenario 7** | Observed Prevalence (%) |  | 3.299 | 5.525 | 9.079 | 14.487 | 30.86 |
|  | Actual Prevalence (%) |  | 5.15 | 8.535 | 13.792 | 21.456 | 42.29 |
|  | Number shutdown | 0% | 144 | 152 | 169 | 195 | 303 |
|  |  | 25% | 0 | 0 | 0 | 0 | 0 |
|  |  | 50% | 0 | 0 | 0 | 0 | 0 |
|  |  | 75% | 256 | 248 | 231 | 205 | 97 |
|  | Number of days of hospital capacity exceeded |  | 0 | 0 | 14 | 26 | 40 |
|  | Number of deaths |  | 3064 | 3755 | 4775 | 6136 | 9607 |

